# Application of the Fluctuation Test to the data of Morbidity and Mortality by COVID-19 in China 2020-2023

**DOI:** 10.1101/2023.11.12.23298174

**Authors:** Anyi Yannire López-Ramírez, María Alejandra Fernández-Ramírez, Hilda Cristina Grassi, Efrén de Jesús Andrades, Jesús Enrique Andrades-Grassi

## Abstract

In this work the Luria and Delbruck Fluctuation Test was applied to the data of Morbidity and Mortality by COVID-19 in China from January 2020 to August 2023. Three types of data were used: es.statista.com, datosmacro.expansion.com and larepublica.co without modification, but trying to avoid and justify the anomalies and inconsistencies observed. The methods originally used to establish the interactions of two populations were evaluated: the viral population with that of its host and the drift of both organisms. Only the fluctuations of the weekly Variance of daily increase of Cases (Morbidity) and of the weekly Variance of daily increase of Deaths (Mortality) were studied. The results showed that the Fluctuation Test is applicable to the selected data from China and other data from India, Japan and South Korea, used as controls. The study was separated into two periods: a first initial period from January 2020 to September 2021 and a second final period from October 2021 to August 2023. Results were obtained for Morbidity and Mortality that relate the fluctuations of the first with the fluctuations of the second. However, it was possible to detect some anomalies and uncertainties that were possibly derived from inconsistencies in the original data. A repeated fluctuation was observed in the boreal winter in January, February and March of each one of the year studied. A clear decrease in fluctuation was detected in that period in 2021 that could be attributed to the strict confinement during the quarantine in China between 2020 and 2021. Massive, extensive and intensive vaccinations failed to completely eliminate the most important fluctuations. In this work we tried to correlate the appearance of some virus variants with the fluctuations. The most relevant results of said correlation are presented. With the results of this work, the animal origin cannot be confirmed nor can the human or laboratory origin of the SARS CoV-2 virus that caused the initial emerging infection, be ruled out. However, it was concluded that this method could be used to search for clues about its origin. One of these keys is the comparison of the result of the first important fluctuation in the boreal winter of 2020 in each of the countries studied as controls: India, Japan and South Korea. The comparison of this result with the first fluctuation of China for that same period could give clues about the origin of the virus.

## Introduction

SARS CoV-2 virus is the etiological agent for COVID-19 outbreak in China **<u>[1]</u>**. This first variant that emerged in a food market in Wuhan, Hubei province in late 2019, produced several derived variants **<u>[2]</u>** as this emerging infection spread worldwide **<u>[3]</u>.** The Chinese government ordered to design, produce and apply several vaccines on the Chinese population from 2021 to 2023 **<u>[4]</u>**. The quarantine in Hubei province started on January 23, 2020. In addition, a policy of strict confinement, control of internal and external mobilization of the population, widespread application of several mandatory diagnostic tests to detect the virus and the disease, among other measures, was ordered in the following years in what was called the Zero COVID-19 Policy **<u>[5]</u>**.

In this work we tried to adapt the Luria and Delbruck Fluctuation Test **<u>[6]</u>** to the data reported on cases and deaths from three sources of information, to verify its applicability, to show and analyze the results and the information that can be produced and to contribute with some aspects such as the Origin of the first variant of the SARS virus CoV-2 **<u>[7]</u>**.

In this work, COVID-19 is considered to be a human infection of Zoonotic origin. What is not yet known is whether or not the virus that caused this emerging infection was previously adapted to human cells. The hypothesis that this work has raised is that if the SARS CoV-2 was not previously adapted to human cells, the first Fluctuation of the boreal winter of 2020 in China should be more greater than the first Fluctuation in that same period of the neighboring countries of China. If both Fluctuations are of the same order, the virus could have been adapted previously to human cells.

## Methods

The Luria and Delbruck Fluctuation Test was adapted according to the method originally used to establish the interaction of two types of populations: The Viral population (SARS CoV-2) and the Host population (human beings from China and other countries).

The veracity of the data was not examined. If in said data there are involuntary or voluntary errors or misconduct, the results, analysis and conclusions of this work will reflect them. Three types of data were used: es.statista.com; datosmacro.expansion.com and larepublica.co without modification, but trying to avoid and justify the anomalies and inconsistencies observed. We proceeded to download the data of the number of confirmed cases and their daily increase (Morbidity). The number of deaths and its daily increase (Mortality) was also downloaded. This data was tabulated in several Excel tables to facilitate its visualization and the calculations of the average and weekly variance of daily increase in Morbidity and Mortality in the years 2020, 2021, 2022 and 2023. Changing the start and end of the days of some of the weeks under study was tested for different data sources, different periods and different countries, to verify whether this shift would produce important changes in the results. The location of the fluctuations was not affected. Furthermore, the values found were of the same order and similar. A discontinuous graphic system was designed to avoid data with anomalies and inconsistencies and to allow the treatment of different data sources, in different periods. These anomalies and inconsistencies were more frequent in the Mortality data than in the Morbidity data. For this reason, discontinuities were reported in some Mortality figures. Moreover, the period in which the full vaccination reached more than 75 per cent and the herd immunity was supposedly hit in China, was considered to separate this study in two periods. Again, taking these factors into account, in this work the study was separated into two large periods, from January 2020 to September 2021 and from October 2021 to August 2023. This last period begins in the month in which more than 75 per cent of the population of China was fully vaccinated. Only fluctuations in the Variance of Cases (Morbidity) and the Variance of Deaths (Mortality) are reported. The Figures, that in most cases were designated as (a) for Morbidity (Variance of cases, Vc, in green color) and designated as (b) for Mortality (Variance of death, Vd, in red color) and the Table, express the Variance values in relation to the Weeks of each year. A maximum total number of Weeks of 52 is considered in each year. In the first period the data from es.statista.com was used and the initial week considered begins on January 23, 2020, so for that year only 50 weeks were considered and the numbering in that first period continued until the end of September 2021. The data from es.statista.com was also used to detect the fluctuation of the boreal winter of 2022. In the second period the data from datosmacro.expansion.com and larepublica.co were used and the numbering begins on October 01 with week number 1 (which is the 40^th^ week of 2021) and continues until the end of August 2023. In some cases, a special analysis is made on a Fluctuation that superimposes another one of lower value that has started earlier in time. The explanation for this result is that probably a smaller and earlier fluctuation in one region of the country studied was quantitatively surpassed by a larger and later one produced in the same or a different region in the same country. Frequently the graphic result obtained resembles a sigmoid curve.

This study does not belong to formal Statistic Sciences. Instead it is an exploratory application of the Fluctuation Test of Luria and Delbruck that evaluates the calculation of Weekly Variance (as VAR.P) of official data reported for COVID-19 cases and death. In this study, the calculation of Weekly Variance (as VAR.P) of cases and death is applied to verify the appearance or not of Fluctuations that would be expressed as a consequence of some events such as quarantine and strict confinement, the appearance of new variants of SARS CoV-2 and vaccination campaign. The calculation of Weekly Variance of the daily increases in cases and deaths from COVID-19 official reported data, were carried out according to VAR.P (number 1,{number 2} …) in the Excel spreadsheet.

## Results

**First Period:** es.statista.com

This period shows the initial fluctuations during the emerging infection caused by SARS CoV-2 in the years 2020 and 2021 in the population of China. In addition, the impact caused by the original variant of the virus in the northern winter of 2020 is shown, which is later used to be compared with the northern winter fluctuation of China in 2021, 2022 and 2023 and also used to be compared with the countries selected as controls in the same period of 2020.

The result with data from es.statista.com of Figure 1(a) shows an important fluctuation in the weekly variance of cases of 22,187,755 between February 10 and 16, 2020. This fluctuation could be contributing to revealing clues for animal or human origin of SARS CoV-2 virus (see below). However, the significant fluctuation in the weekly variance of deaths occurs nine weeks later with a value of 203,609 (see Figure 1(b)). An anomalous delayed result similar to this occurred between November 2022 and January 2023 (see Figures 6 and 7).

**Figure 1(a).**
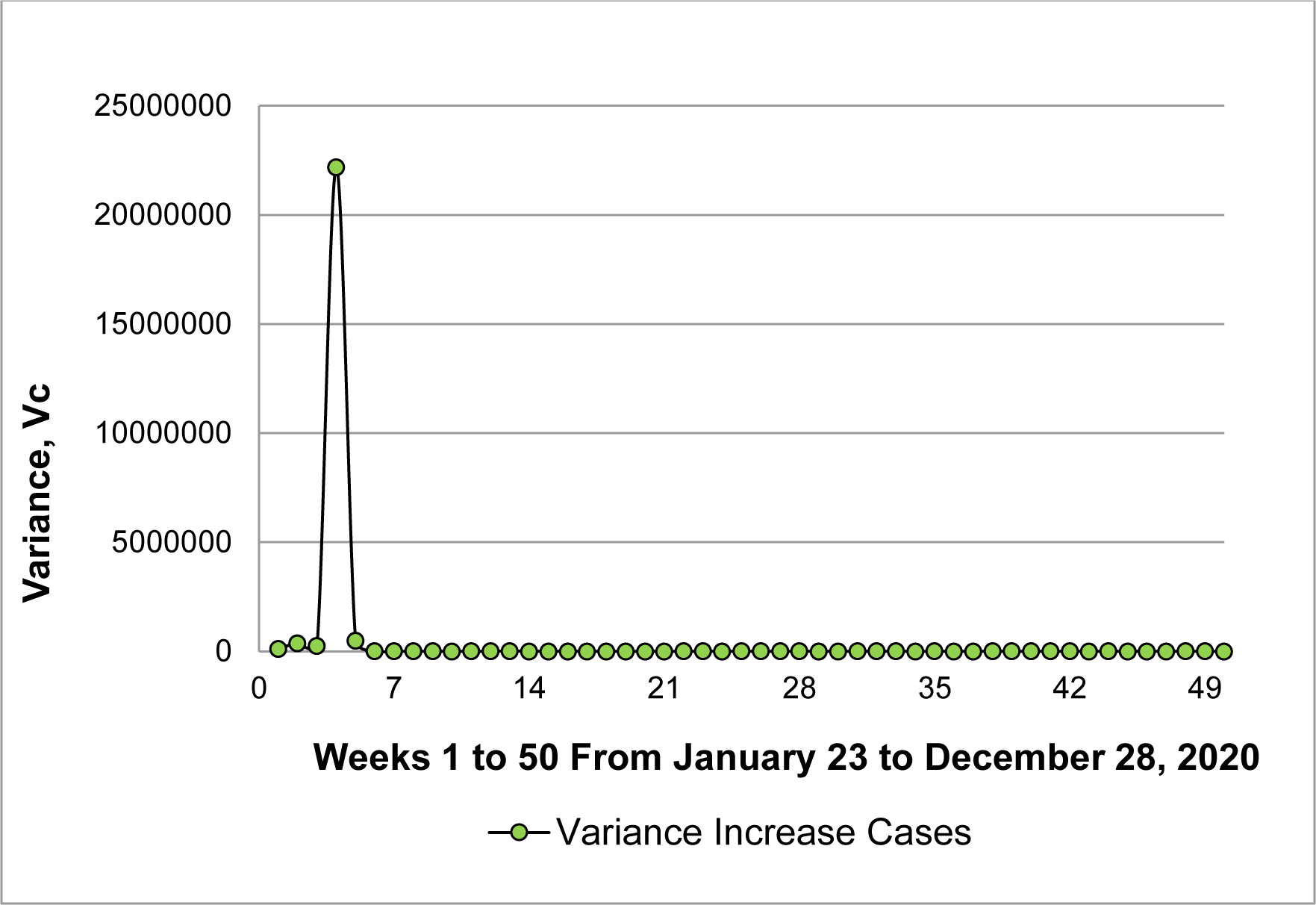
Variance of weekly increase in cases per day based on the 50 weeks from January 23 to December 28, 2020.

**Figure 1(b).**
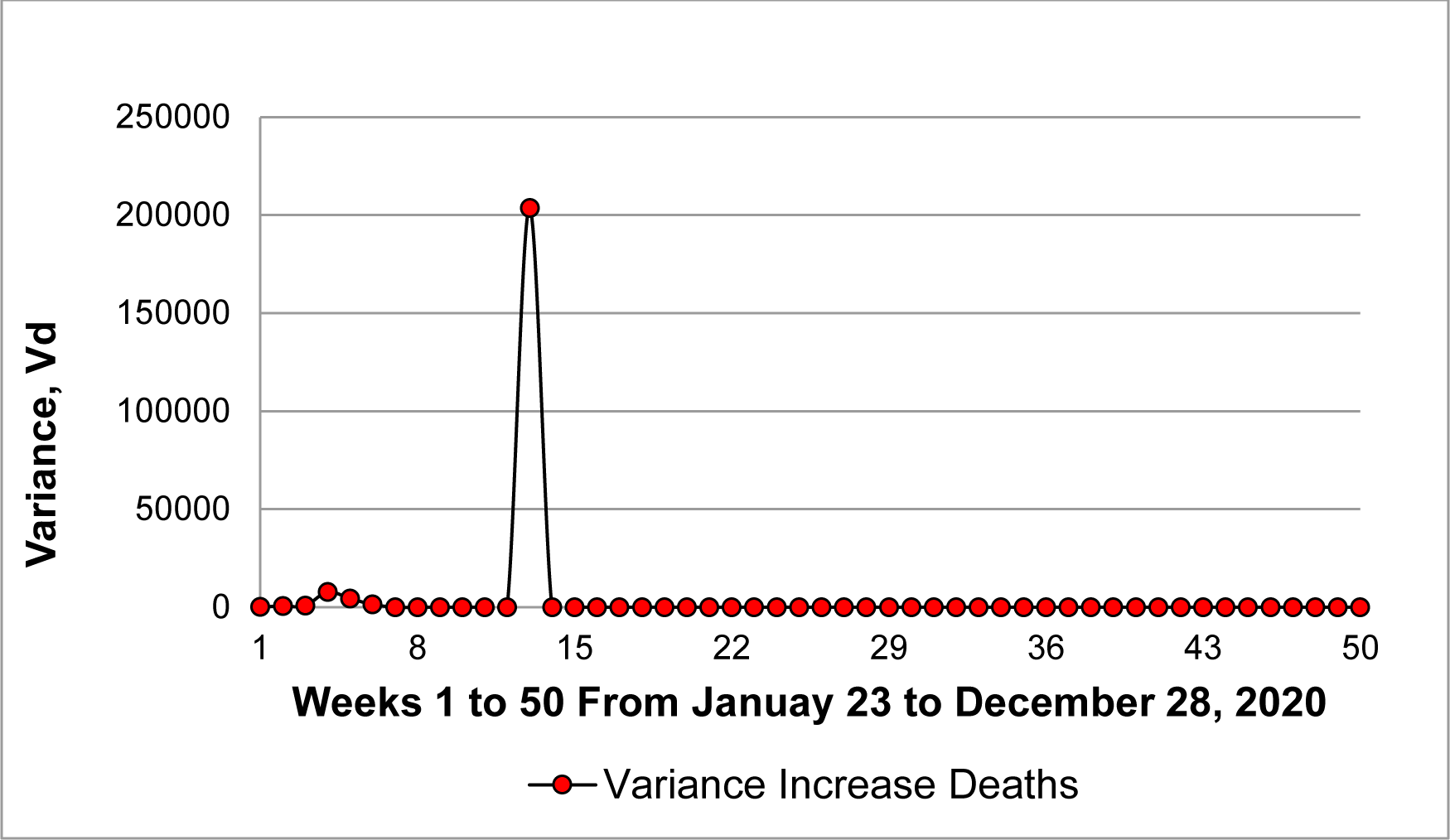
Variance of weekly increase in deaths per day based on the 50 weeks from January 23 to December 28, 2020.

In the year 2021 (weeks 3 to 39), it was observed that the highest values in the fluctuations of the weekly variance of cases (Figure 2(a)) and the weekly variance of deaths (Figure 2(b)) were 6,938 (week 6) and 0.82 (week 8), respectively. These low values would be explained due to the strict confinement of the quarantine in China.

**Figure 2(a).**
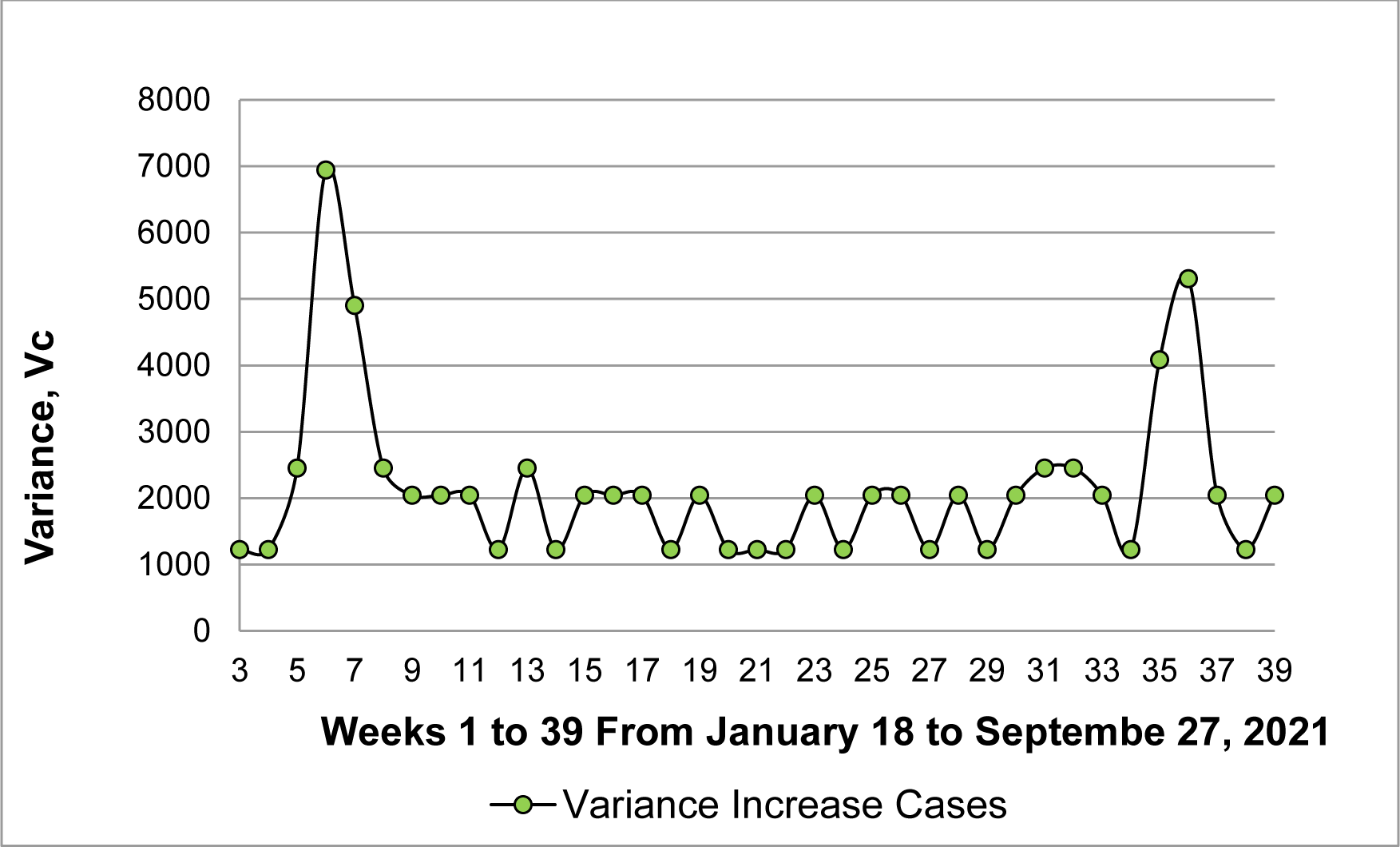
Variance of weekly increase in cases per day based on the weeks 3 to 39 from January 18 to September 27, 2021.

**Figure 2(b).**
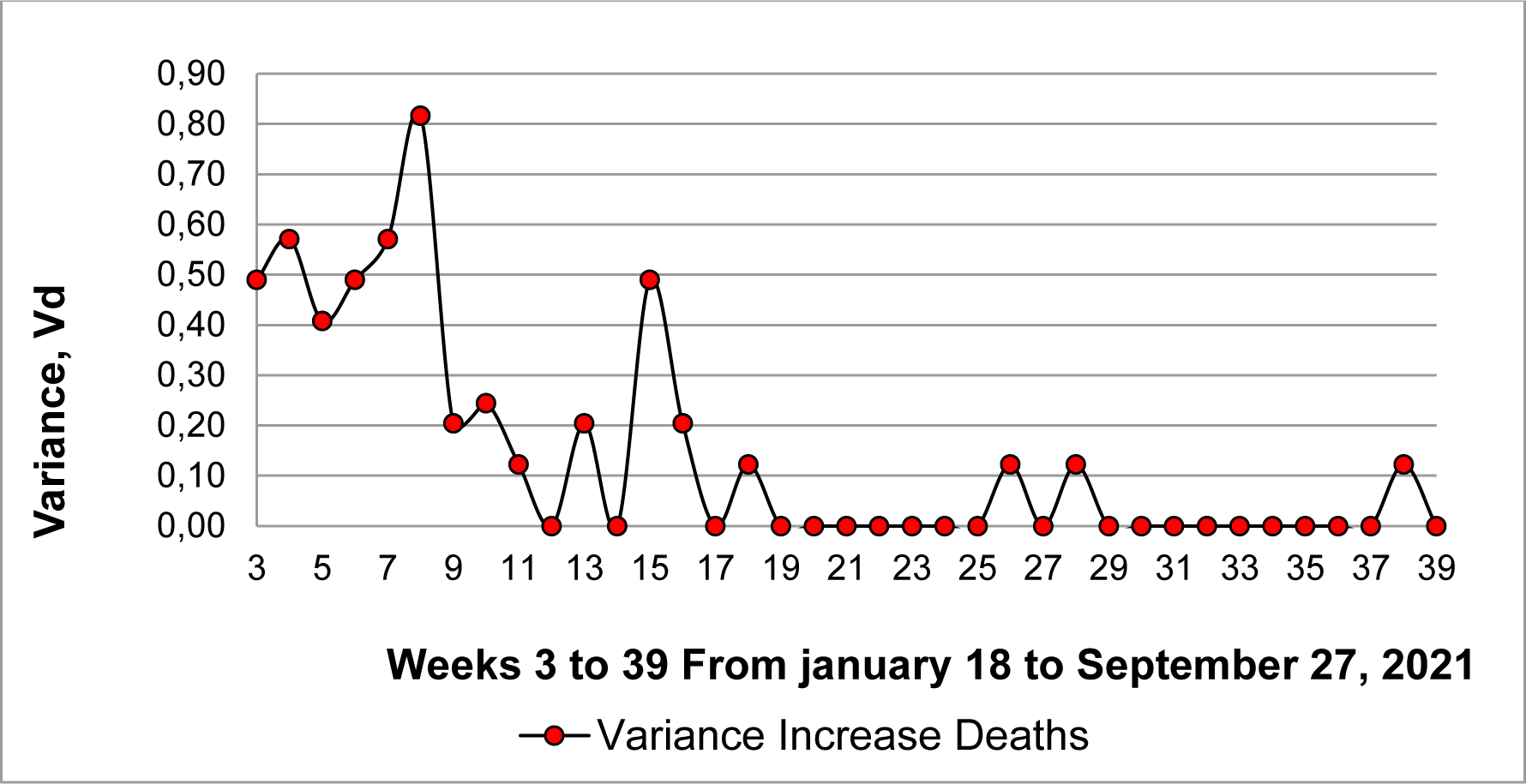
Variance of weekly increase in deaths per day based on the weeks 3 to 39 from January 18 to September 27, 2021.

The Figure 3(a) again shows a fluctuation in the northern winter of 2022 using data from es.statista.com. A similar result was observed in this period using data from datosmacro.expansion.com and larepublica.co, with some differences (see below). Again, these first 15 weeks of the northern winter period are important for a fluctuation to occur. This result was confirmed for death in Figure 3(b). The results shown in Figures 3(a) and 3(b) would also confirm the possible role of an emerging variant (Omicron) of the virus after November 2021 **<u>[8]</u>** and the possible failure of the vaccination campaign in China after October 2021.

**Figure 3(a).**
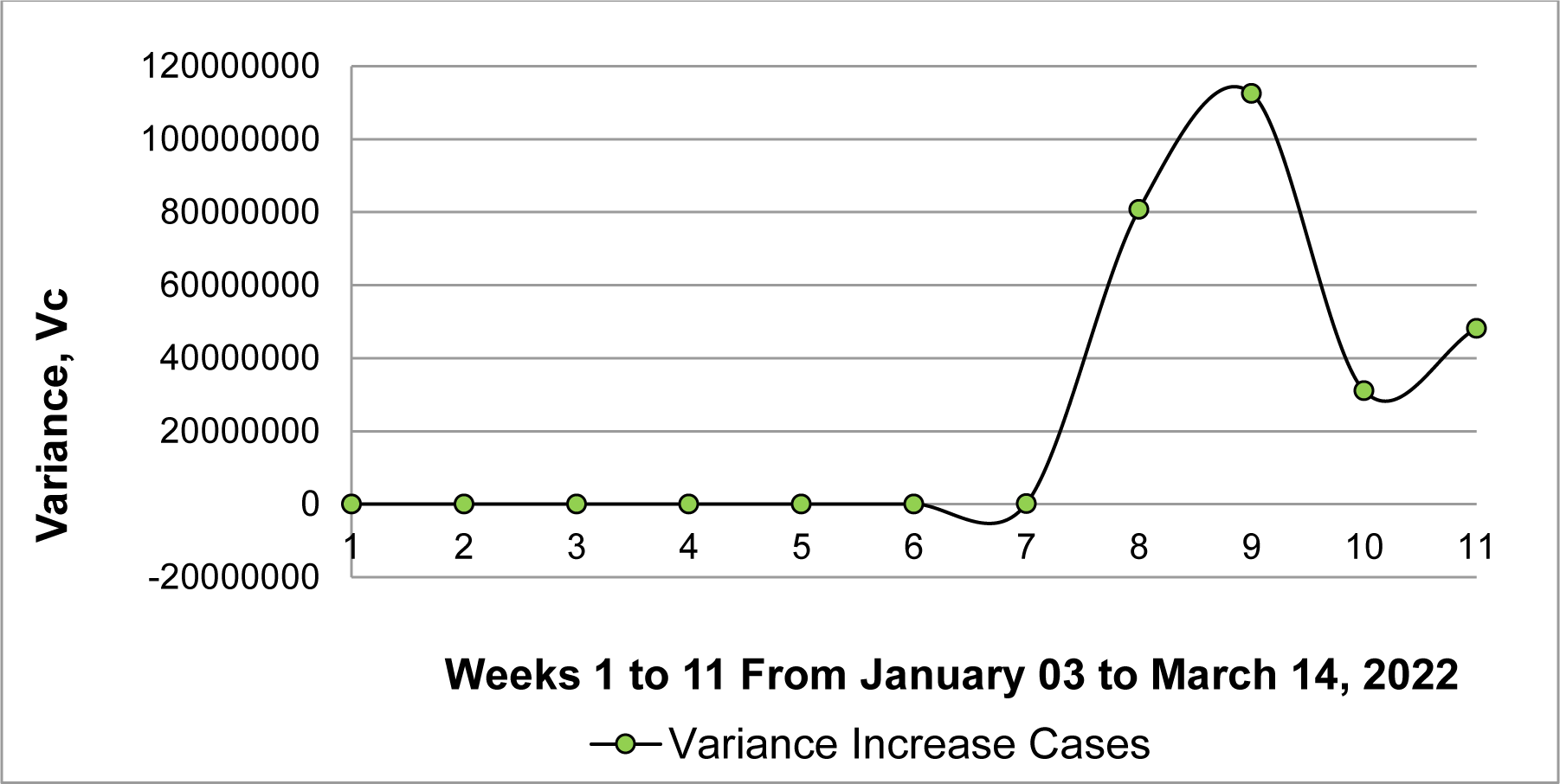
Variance of weekly increase in cases per day based on the 11 weeks from January 03 to March 14, 2022. Y-axis negative value is due only to the tendency not to the points.

**Figure 3(b).**
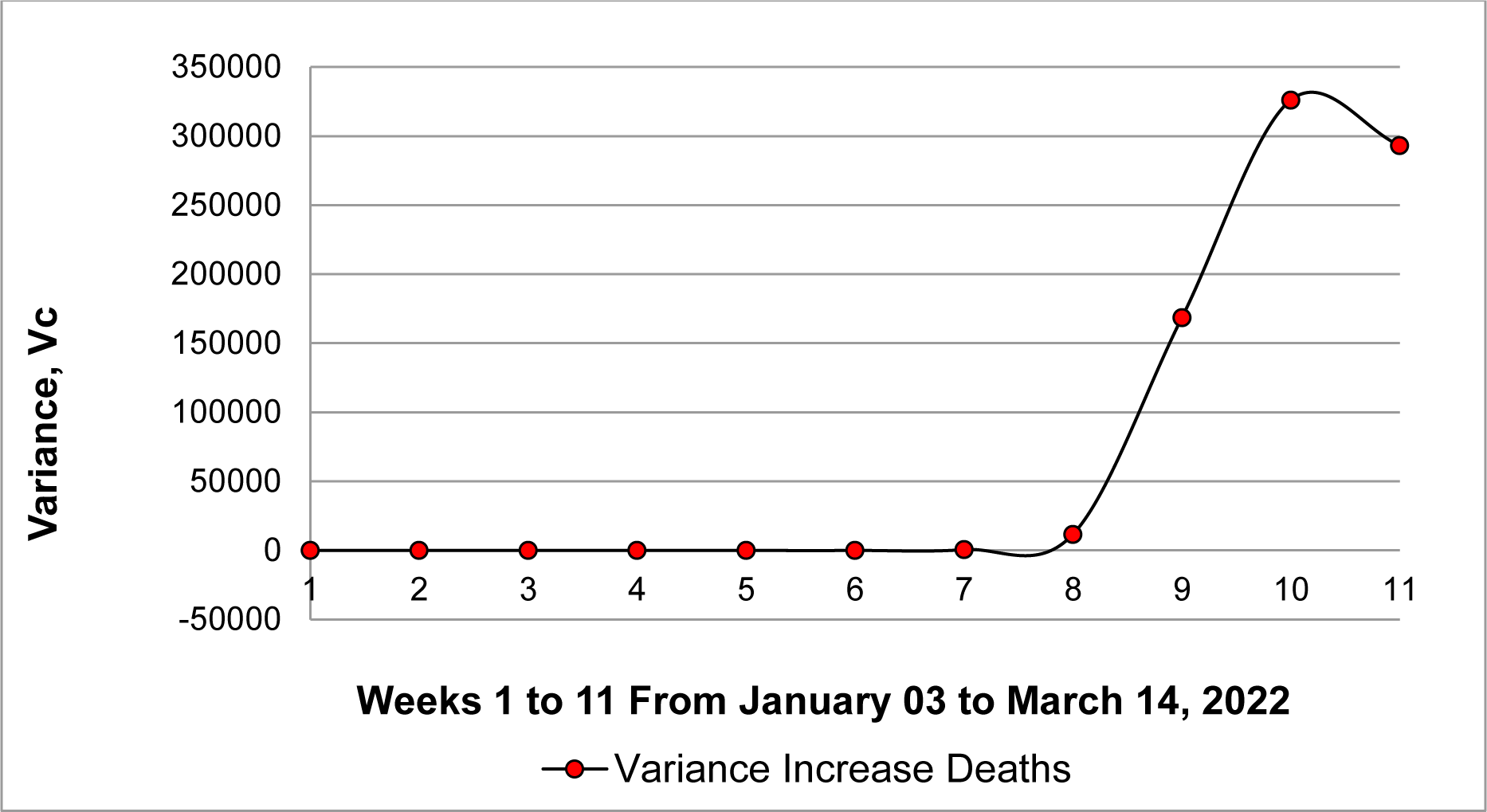
Variance of weekly increase in deaths per day based on the 11 weeks from January 03 to March 14, 2022. Y-axis negative value is due only to the tendency not to the points.

The Figure 3(b) shows an important fluctuation in the weekly variance of deaths of 326,177 from March 07 to 13, 2022.

**Second Period:** datosmacro.expansion.com and larepublica.co

This period shows the final fluctuations from October 2021 to August 2023. This last period begins in the month in which more than 75 per cent of the population of China was fully vaccinated. Furthermore, infections caused by Omicron, one of the most important variants of SARS CoV-2, occur in this period in China. In addition, the impact caused by the original variant of the virus in the northern winter of 2020 is shown, which is later used to compare with the northern winter fluctuation of China in 2021, 2022 and 2023.

In this period the variance of weekly increase of death is considered first because the weekly variance increase of cases is subject to special analysis (see below).

Two significant fluctuations occur around week 22 and week 52 (Figure 4(a)), its variance values are 4,367 and 2,981, respectively. In addition, small fluctuations of the order of 500 are observed between weeks 24 and 32.

**Figure 4(a).**
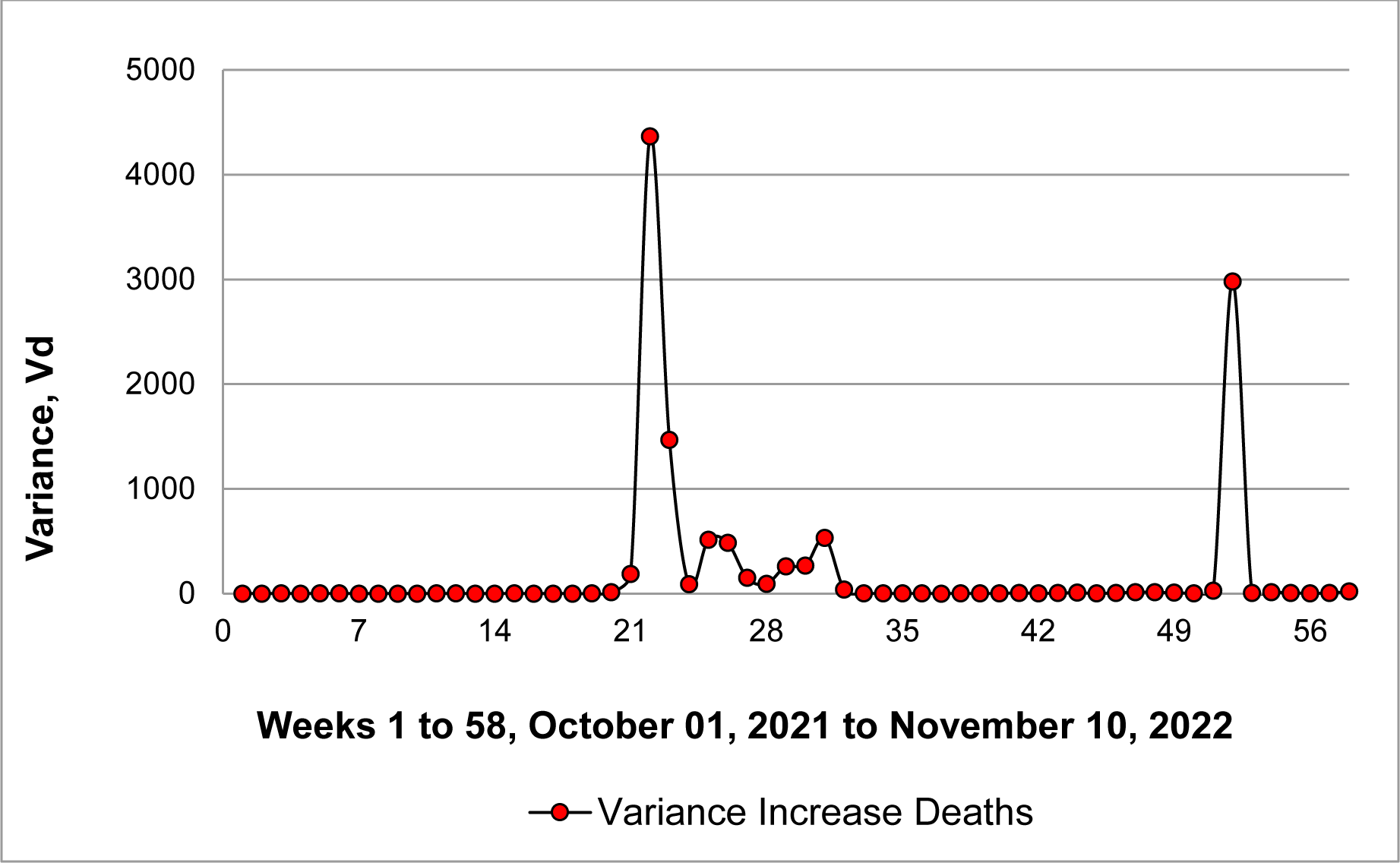
Variance of weekly increase in deaths per day based on the 58 weeks from October 01, 2021 to November 10, 2022.

The result of Figure 4(b) in the same period of Figure 4(a) shows an apparent single important fluctuation whose maximum level occurs in week 22 with a variance value of 476,228,501.

**Figure 4(b).**
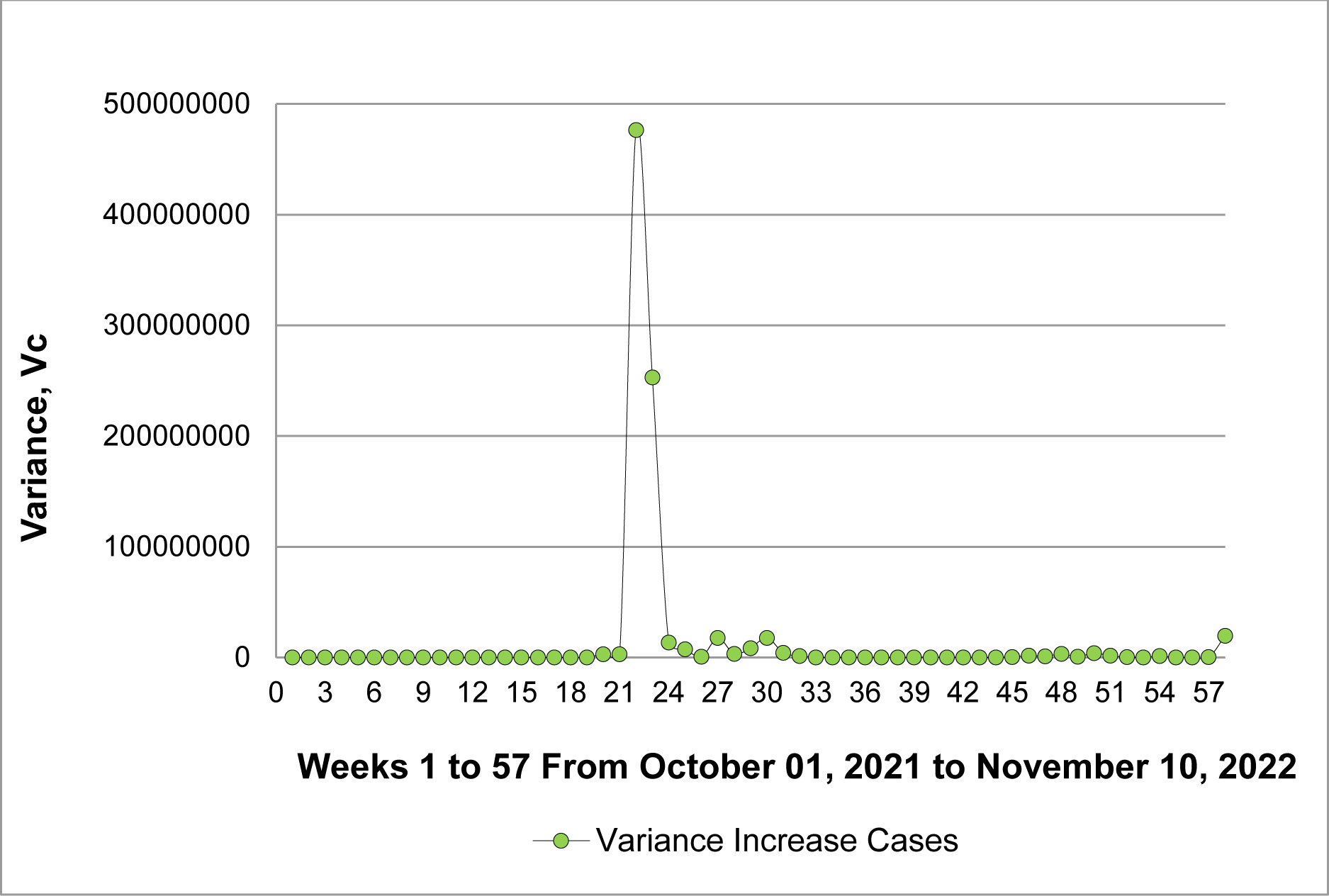
Variance of weekly increase in cases per day based on the 58 weeks from October 01, 2021 to November 10, 2022.

The scale imposed on the Y axis of Figure 4(b) does not allow the fluctuations shown in Figure 5 (weeks 40 to 55) to be appreciated. These fluctuations in the weekly variance of cases possibly lead to the fluctuation of the weekly variance of deaths in week 52 of Figure 4(a), as it happens in week 22 (see comparatively Figures 4(a) and 4(b)). These results demonstrate that fluctuations in the weekly variance of cases should drive the observed fluctuations in the weekly variance of deaths in the same or consecutive weeks. However, the large asymmetrical fluctuation in the weekly variance of cases shown in Figure 6 (weeks 55 to 70) do not produce an immediate and proportional fluctuation in the weekly variance of deaths in these weeks (see Figure 7). This Figure 7 shows an important delayed fluctuation eight weeks later than the fluctuation in variance ofcases on week 60. A similar delay was observed in the first period (see Figures 1(a) and 1(b)).

**Figure 5.**
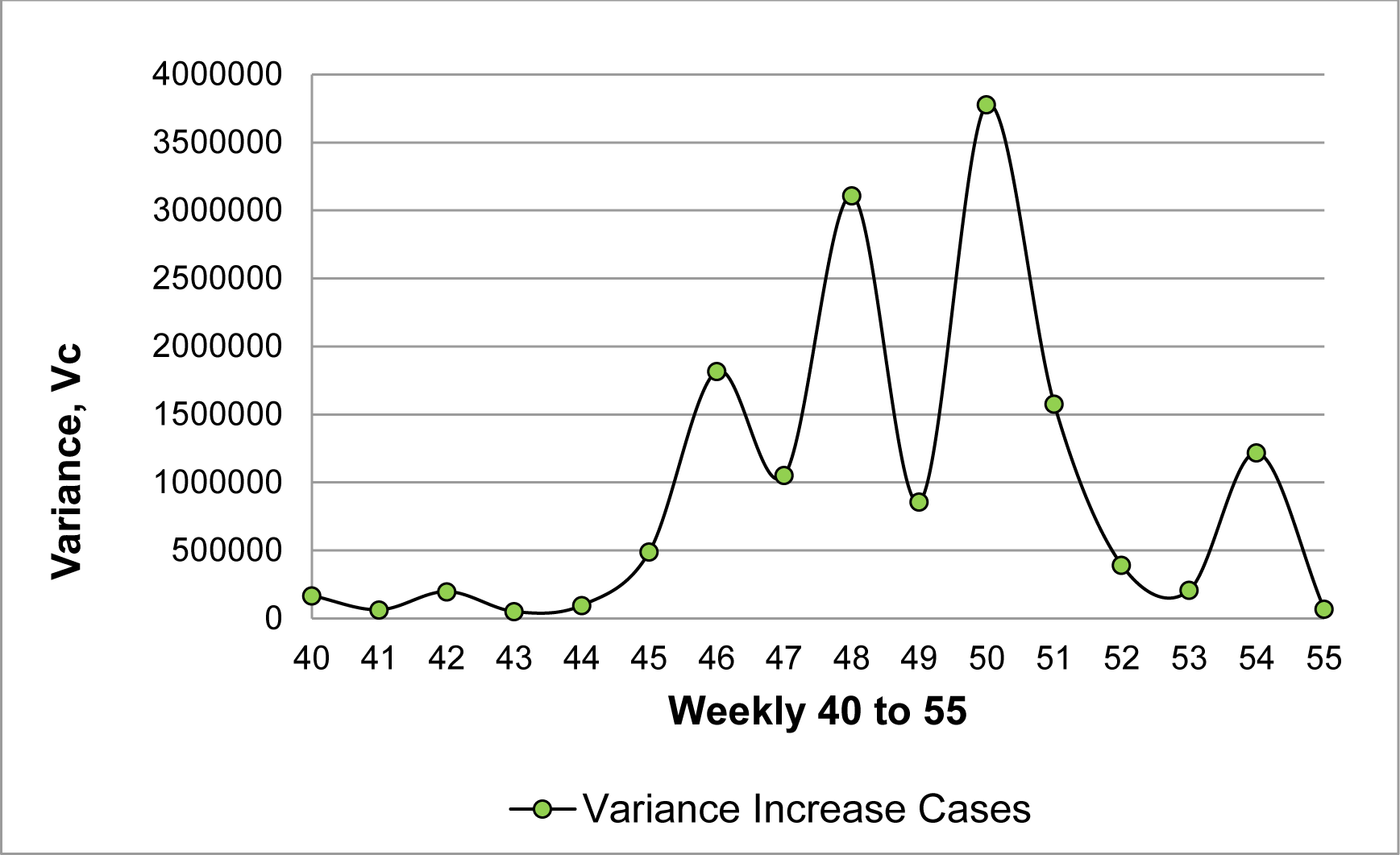
Variance of weekly increase in cases per day based on the 15 weeks from July 01, 2022 to October 14, 2022.

**Figure 6.**
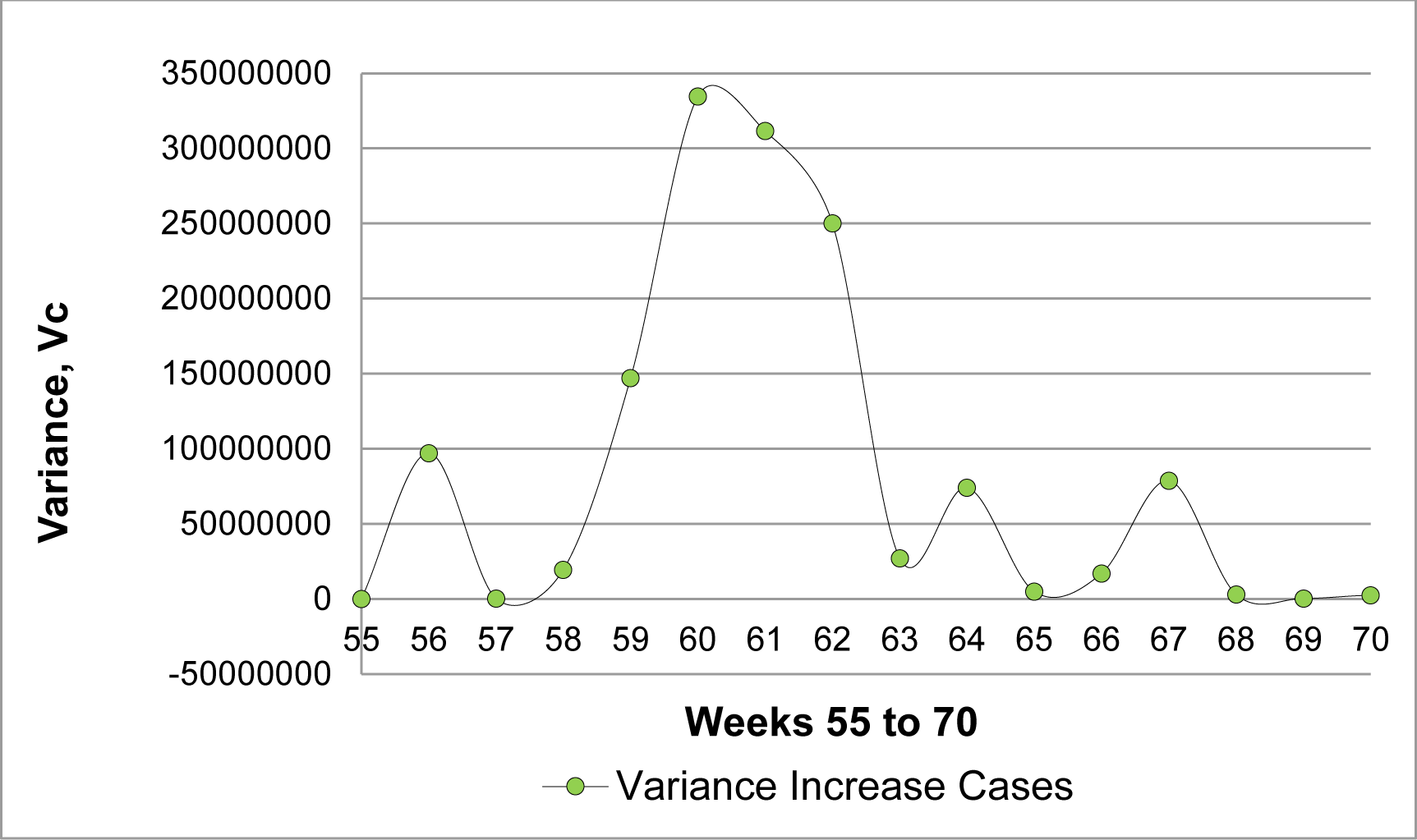
Variance of weekly increase in cases per day based on the 16 weeks from October 14, 2022 to February 02, 2023. Y-axis negative value is due only to the tendency not to the points.

**Figure 7.**
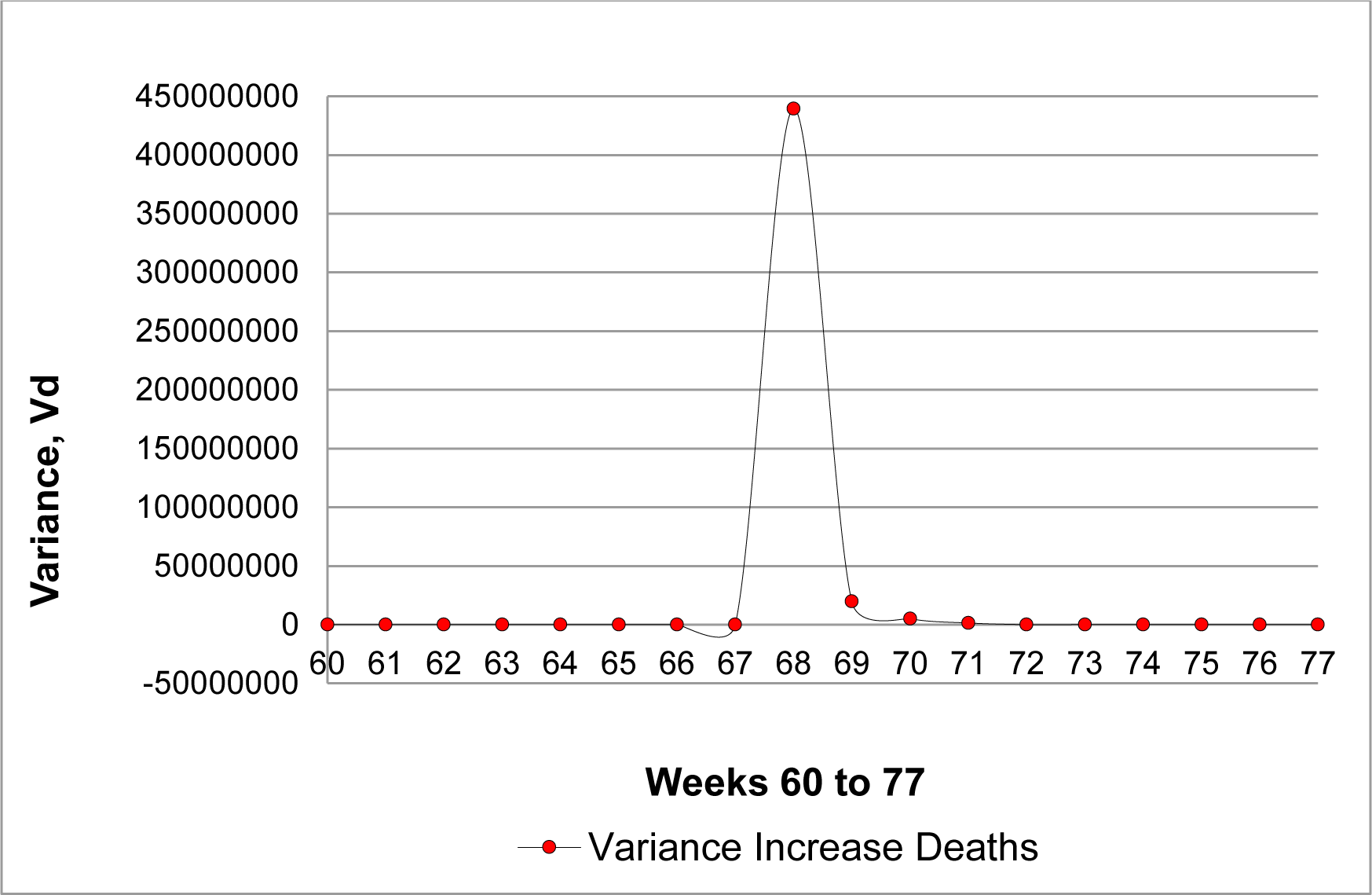
Variance of weekly increase in deaths per day based on the 18 weeks from November 18, 2022 to March 23, 2023. Y-axis negative value is due only to the tendency not to the points.

In Table 1 the fluctuations of the weekly variance of COVID-19 cases in the Chinese population are observed for the beginning of winter in the northern hemisphere in years 2020, 2021, 2022 and 2023, which also coincides with one of the most important festivities in China: The Chinese New Year, this is also an important time for international tourism and the internal and external mobilization of citizens residing in that country.

**Table 1.** Comparison of the most important fluctuations in the boreal winter period for the months January, February and March 2020, 2021, 2022 and 2023.

| Year | Weekly Period | es.statista.com<br>Fluctuation Value in<br>Vc and Vd | datosmacro.expansion.com<br>and larepublica.co<br>Fluctuation Value in Vc and Vd |
| --- | --- | --- | --- |
| 2020 | February 10 to 16 | 22,187,755 and<br>7,770* | - |
|  | February 13 to 19 | - | 22,734,585 |
|  | February 20 to 26 | - | 5,059* |
|  | April 13 to 19 | 203,609** | - |
| 2021 | January 18 to 24 | 6,938 | - |
|  | February 15 to 21 | 0.82 | - |
|  | January 21 to 27 | - | 2,885 |
|  | January 28 to<br>February 03 | - | 2.78 |
| 2022 | February 25 to<br>March 03 | - | 476,228,501 |
|  | March 04 to 10 | - | 1,466 |
|  | February 28 to<br>March 06 | 112,562,040 | - |
|  | March 07 to 13 | 326,177 | - |
| 2023 | January 06 to 12 | - | 78,814,952 |
|  | January 13 to 19 | - | 439,431,344 |

**Note:** The red color represents the weekly variance of daily increases in deaths and the green color represents the weekly variance of daily increases in confirmed Covid-19 cases in the Chinese population. \* These two values derived from two different data are similar, even though they correspond to different weeks of the same period in February 2020. \*\*The value of 203,609 corresponds to a nine-week delay of the es.statista.com data for that period.

In every year a significant increase is observed in a few weeks of that period, however, the maximum values reflect the differences in human activity in each of those years. In 2020, the weekly variance of cases exceeded 22 million. This period coincides with the initial outbreak of the pandemic, which could point to the possible origin of the virus.

By 2021, under the effect of strict confinement, the maximum value of the weekly variance of cases decreased to almost 7,000 and then increased to more than 476 millions in February-March 2022, and to a maximum of 78 million in January 2023. In these last two years, the Chinese population was partially released from confinement. These two increases occurred despite the fact that for the last two years, the vaccination process had progressed significantly. Table 1 shows the values of fluctuations in the boreal winter of first and second period using es.statista.com, datosmacro.expansion.com and larepublica.co

The results in the Table 1 would confirm the validity of applying the fluctuation test to these three source of data in a period in which both, weekly variance of daily increase of cases and death, show important fluctuations. It is observed that in the boreal winter period in the months January, February, and March of the years 2020, 2021, 2022 and 2023, there are relevant increases in the fluctuations of the weekly variance of cases and deaths.

**Clues for the possible origin of SARS CoV-2:** larepublica.co

The results for variance of cases and variance of death in the boreal winter of 2020 in South Korea (Figures 8(a) and 8(b)), Japan (Figures 9(a) and 9(b)) and India (Figures 10(a) and 10(b)) are clearly lower in several order of magnitude than the Fluctuations values of 22,187,755 and 203,609 obtained for variance of cases and variance of death, respectively, in the same period in China.

**Figure 8(a).**
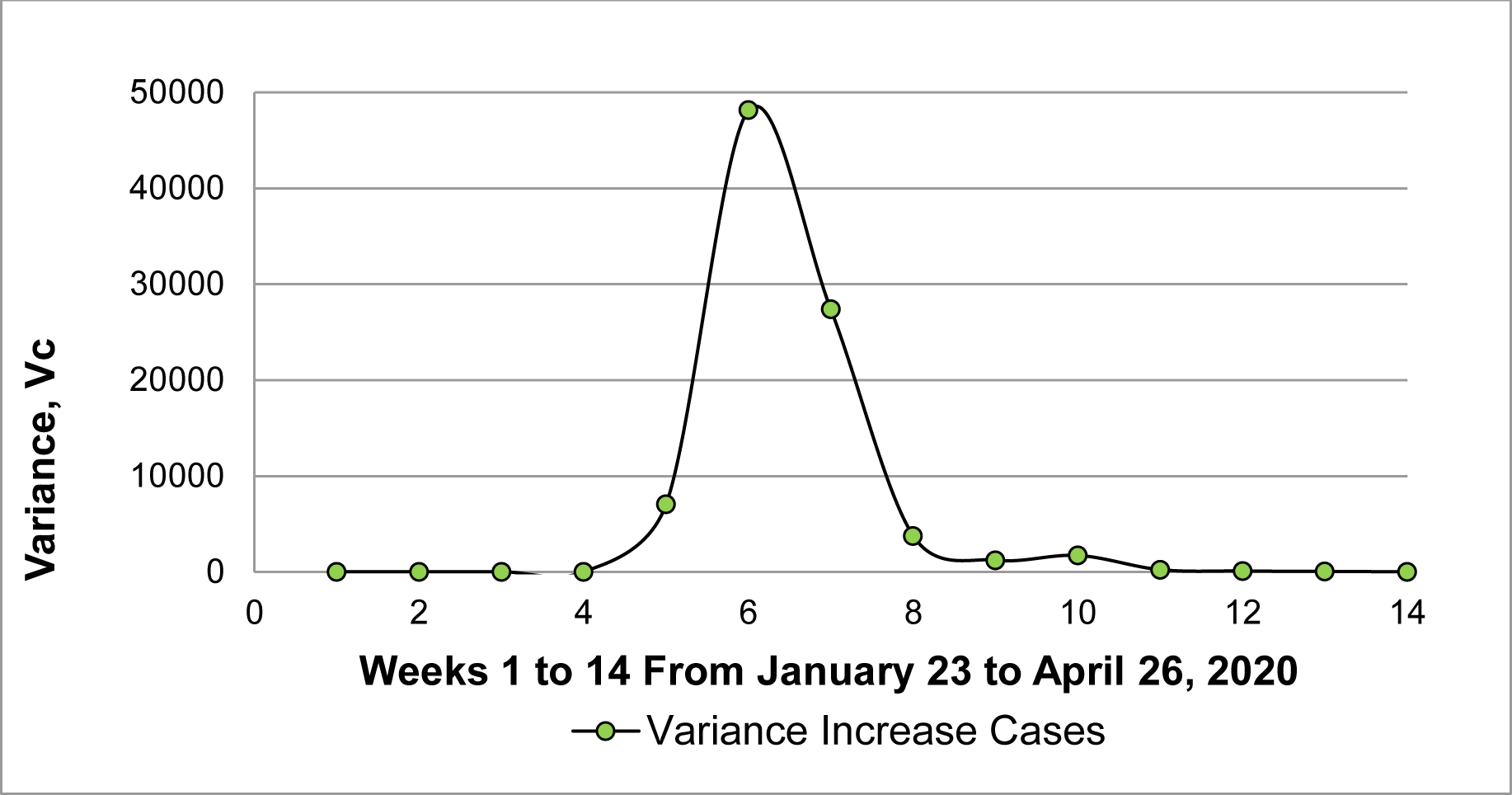
**South Korea:** Variance of weekly increase in cases per day based on the 14 weeks from January 23 to April 26, 2020.

**Figure 8(b).**
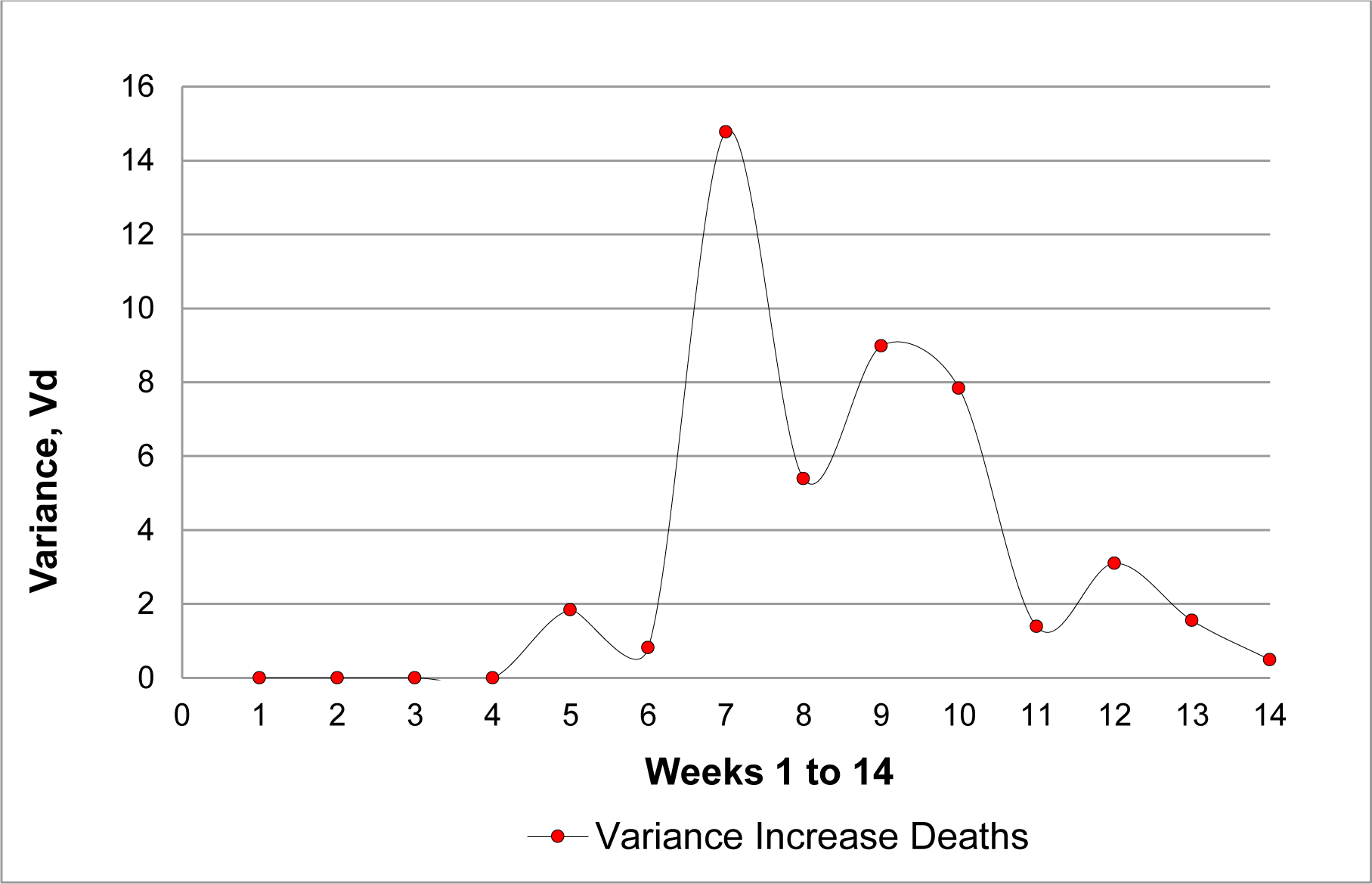
**South Korea:** Variance of weekly increase in deaths per day based on the 14 weeks from January 23, 2020 to April 26, 2020.

**Figure 9(a).**
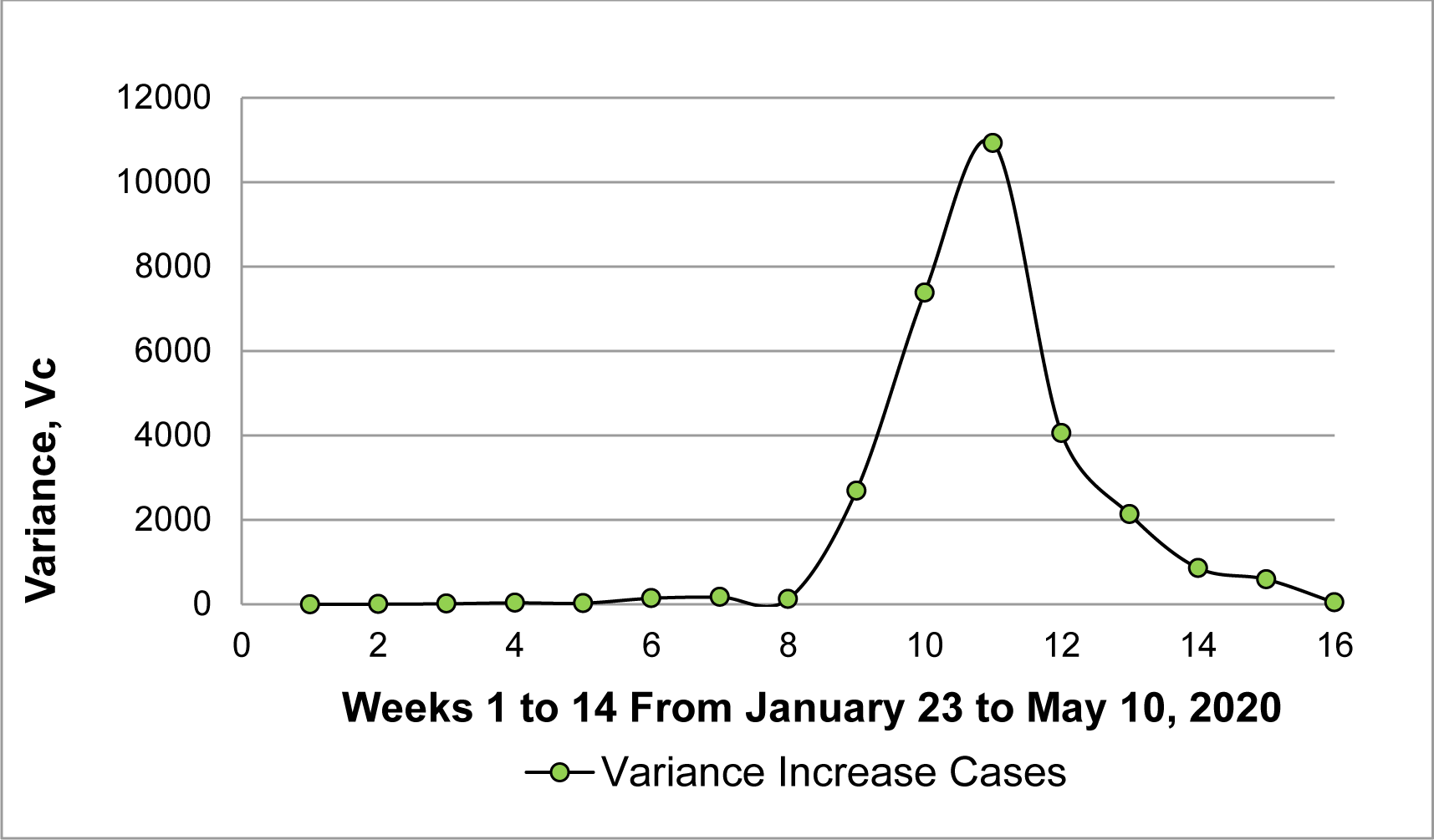
**Japan:** Variance of weekly increase in cases per day based on the 14 weeks from January 23 to May 10, 2020.

**Figure 9(b).**
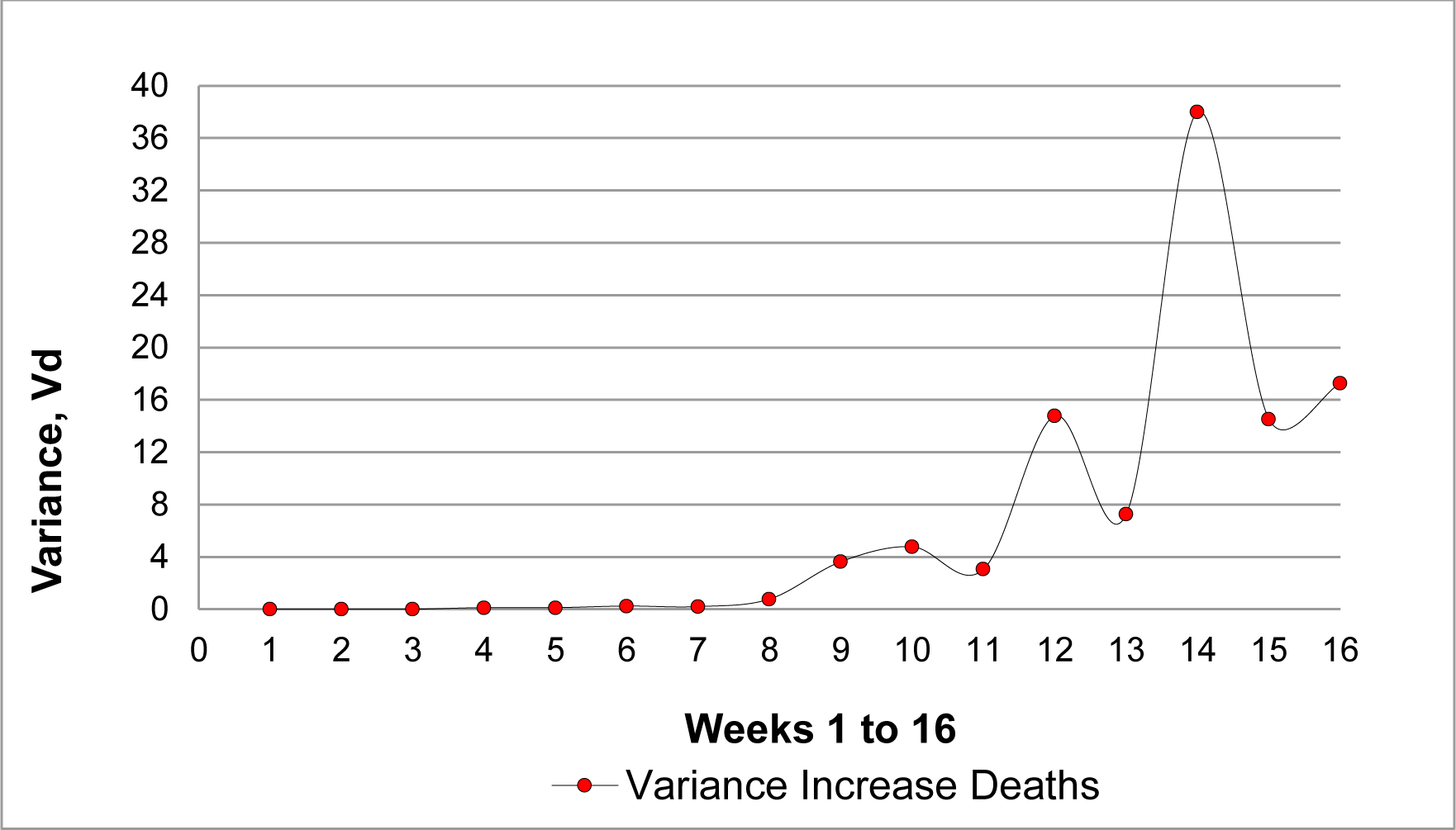
**Japan:** Variance of weekly increase in deaths per day based on the 16 weeks from January 23, 2020 to May 10, 2020.

**Figure 10(a).**
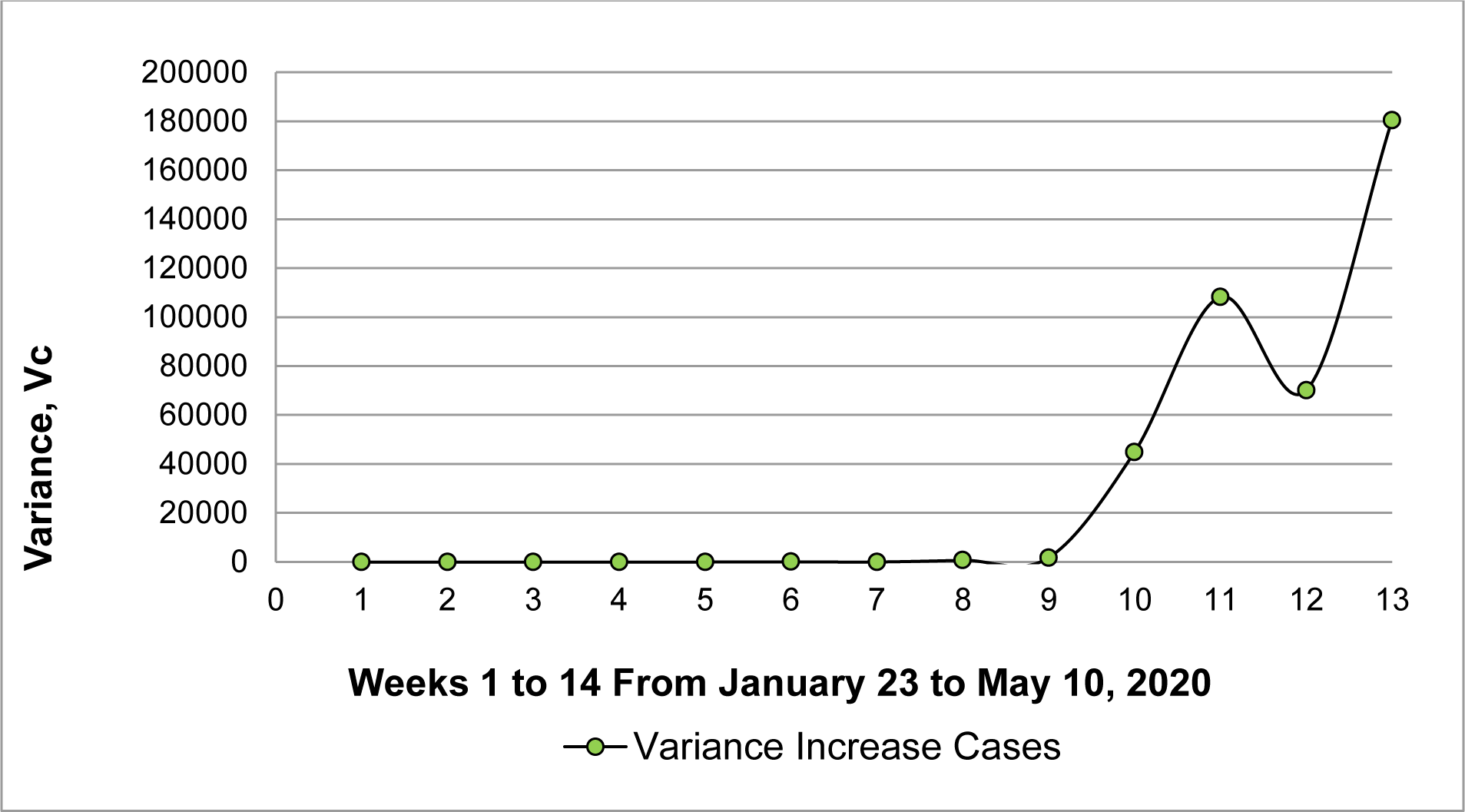
**India:** Variance of weekly increase in cases per day based on the 13 weeks from January 23 to April 19, 2020.

**Figure 10(b).**
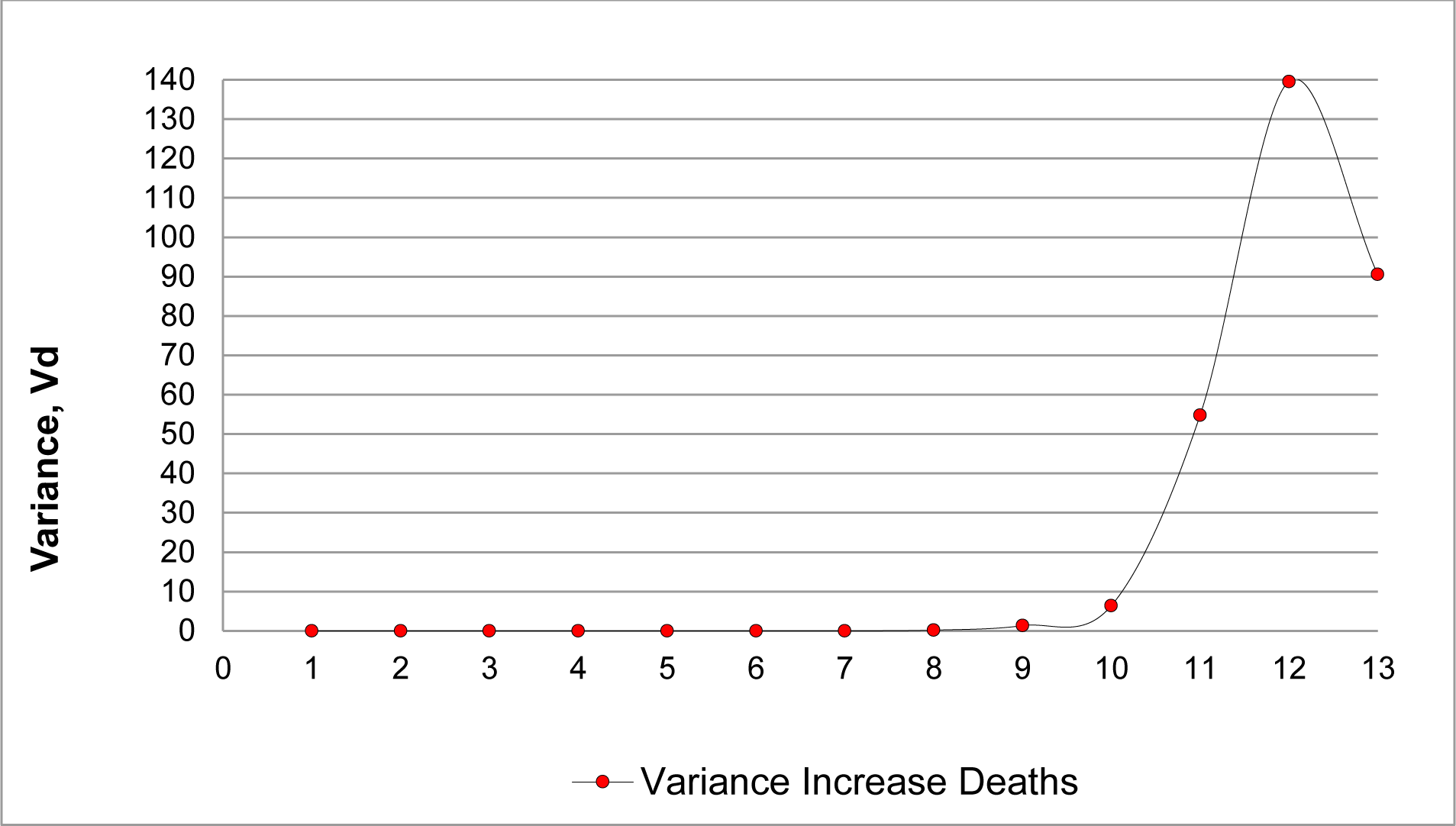
**India:** Variance of weekly increase in deaths per day based on the 16 weeks from January 23, 2020 to April 19, 2020.

Relative low values of variance of cases and variance of deaths for South Korea (week 6, V_c_=48,149 and week 7, V_d_=14.78), for Japan (week 11, V_c_=10,929 and week 14, V_d_=38.00) and for India (week 11, V_c_=108,281 and week 12, V_d_=139.55), seem to point out that high value of variance of cases and variance of death in the northern winter of China in 2020, would confirm the high impact of COVID-19 in the human population of this country, target of the outbreak of SARS Cov-2 infection. The high impact of the change of host for the virus may explain the difference of Fluctuations between China and neighboring countries.

Early fluctuations of India in the boreal winter of 2020 (see Figures 10(a) and 10(b)) show an additional increase in the values of variance of cases and variance of deaths in week 13. This increase belong to a second, later and much larger fluctuation that reached its maximum by the end of spring of 2020 (V_c_ more than 487 million in week 25 and V_d_ more than 331 thousand in week 22, results not shown). We perform a special analysis to explain the effect of a fluctuation that superimposes another one of lower value that has started earlier in time in the same country. Frequently graphic result obtained resembles a Sigmoid curve in several data analysis that are not shown.

## Conclusions

The data used in this work absolutely does not include the participation or opinion of the authors. It is declared that due to this, results, analysis and conclusions depend strictly on the veracity of said data.

The results showed that the Fluctuation Test is applicable to the selected data from China and other data from India, Japan and South Korea. The methods originally used to establish the interactions of two populations were evaluated: the viral population with that of its host and the drift of both organisms. The study was separated into two periods: a first initial period from January 2020 to September 2021 and a second final period from October 2021 to August 2023.A repeated fluctuation was presented in the boreal winter in January, February and March of each one of the year studied. A clear decrease in Fluctuation was detected in that period in 2021 that could be attributed to the strict confinement during the quarantine in China between 2020 and 2021.Massive, extensive and intensive vaccinations failed to completely eliminate the most important fluctuations.In this work we tried to correlate the appearance of some virus variants with the fluctuations. The most relevant result is observed between the end of 2021 and the beginning of 2022, in the period in which the Omicron variant was activated. With the results of this work, the animal origin cannot be confirmed nor can the human origin of the SARS CoV-2 that caused the initial emerging infection, be ruled out. However, it was concluded that this method could be used to search for clues about its origin. In this work one of these keys is the comparison of the result of the first important fluctuation in the boreal winter of 2020 in each of the countries studied as controls: India, Japan and South Korea. The comparison of this result with the first fluctuation of China for that same period gives indications that the origin of the virus may be from an animal, without pre adaptation to Human Cells, but this work does not completely rule out the possibility that this virus is of human or laboratory origin.

## Data Availability

All data produced in the present study are available upon reasonable request to the authors

## Acknowledgements

This work was financed by the Authors. Authors declares no Competing nor Conflict of interests.

## Author Contribution

HCG, EDJA and JEA-G: Conceived and designed the analysis. Contibuted analysis tools.

AYL-R and MAF-R and EDJA: Collected the data and performed the analysis.

AYL-R, MAF-R and EDJA: Drafting the manuscript and wrote final version.

